# Clinical Applications and Limitations of Large Language Models in Nephrology: A Systematic Review

**DOI:** 10.1101/2024.10.30.24316199

**Authors:** Zoe Unger, Shelly Soffer, Orly Efros, Lili Chan, Eyal Klang, Girish N Nadkarni

**Affiliations:** First Faculty of Medicine, Charles University, Prague, Czech Republic; Institute of Hematology, Davidoff Cancer Center, Rabin Medical Center, Petah-Tikva, Israel; School of Medicine, Tel Aviv University, Tel Aviv, Israel; National Hemophilia Center and Thrombosis Institute, Sheba Medical Center, Ramat Gan, Israel; The Division of Data-Driven and Digital Medicine (D3M), Icahn School of Medicine at Mount Sinai, New York, New York, USA; The Charles Bronfman Institute of Personalized Medicine, Icahn School of Medicine at Mount Sinai, New York, New York, USA; The Barbara T Murphy Division of Nephrology, Icahn School of Medicine at Mount Sinai, New York, New York

## Abstract

**Background:** Large Language Models (LLMs) are emerging as promising tools in healthcare. This systematic review examines LLMs’ potential applications in nephrology, highlighting their benefits and limitations.

**Methods:** We conducted a literature search in PubMed and Web of Science, selecting studies based on Preferred Reporting Items for Systematic Reviews and Meta-Analyses (PRISMA) guidelines. The review focuses on the latest advancements of LLMs in nephrology from 2020 to 2024. PROSPERO registration number: CRD42024550169.

**Results:** Fourteen studies met the inclusion criteria and were categorized into five key areas of nephrology: Streamlining workflow, disease prediction and prognosis, laboratory data interpretation and management, renal dietary management, and patient education. LLMs showed high performance in various clinical tasks, including managing continuous renal replacement therapy (CRRT) alarms (GPT-4 accuracy 90-94%) for reducing intensive care unit (ICU) alarm fatigue, and predicting chronic kidney diseases (CKD) progression (improved positive predictive value from 6.7% to 20.9%). In patient education, GPT-4 excelled at simplifying medical information by reducing readability complexity, and accurately translating kidney transplant resources. Gemini provided the most accurate responses to frequently asked questions (FAQs) about CKD.

**Conclusions:** While the incorporation of LLMs in nephrology shows promise across various levels of patient care, their broad implementation is still premature. Further research is required to validate these tools in terms of accuracy, rare and critical conditions, and real-world performance.

## Introduction

Large language models (LLMs), such as ChatGPT^1^, are advanced AI models designed to generate human-like text^2^. These models have already shown potential across various medical specialties^3–11^. The complex nature of kidney diseases and their treatment may enable LLM technology to improve clinical management.

Multimodal LLMs allow for effective interpretation of complex data, including visual data through imaging^12^. They are also capable of tailoring treatments by accessing evidence-based scientific literature^13^. Furthermore, LLMs can possibly automate routine tasks, such as documenting medical records, analyzing laboratory tests, and reviewing different imaging modalities^14–16^. This automation may allow doctors to focus more on providing patient-centered care, ultimately leading to better outcomes in nephrology practice.

In this review, we aim to show diverse clinical applications of LLM in the field of nephrology. Our review outlines the capabilities and limitations of LLM in kidney disease management.

## Overview of AI modalities (Figure 2)

*Artificial Intelligence (AI)* aims to train a computer to perform tasks usually requiring human cognition. AI is a general term referring to a broad range of models^17^.

**Figure 1:**
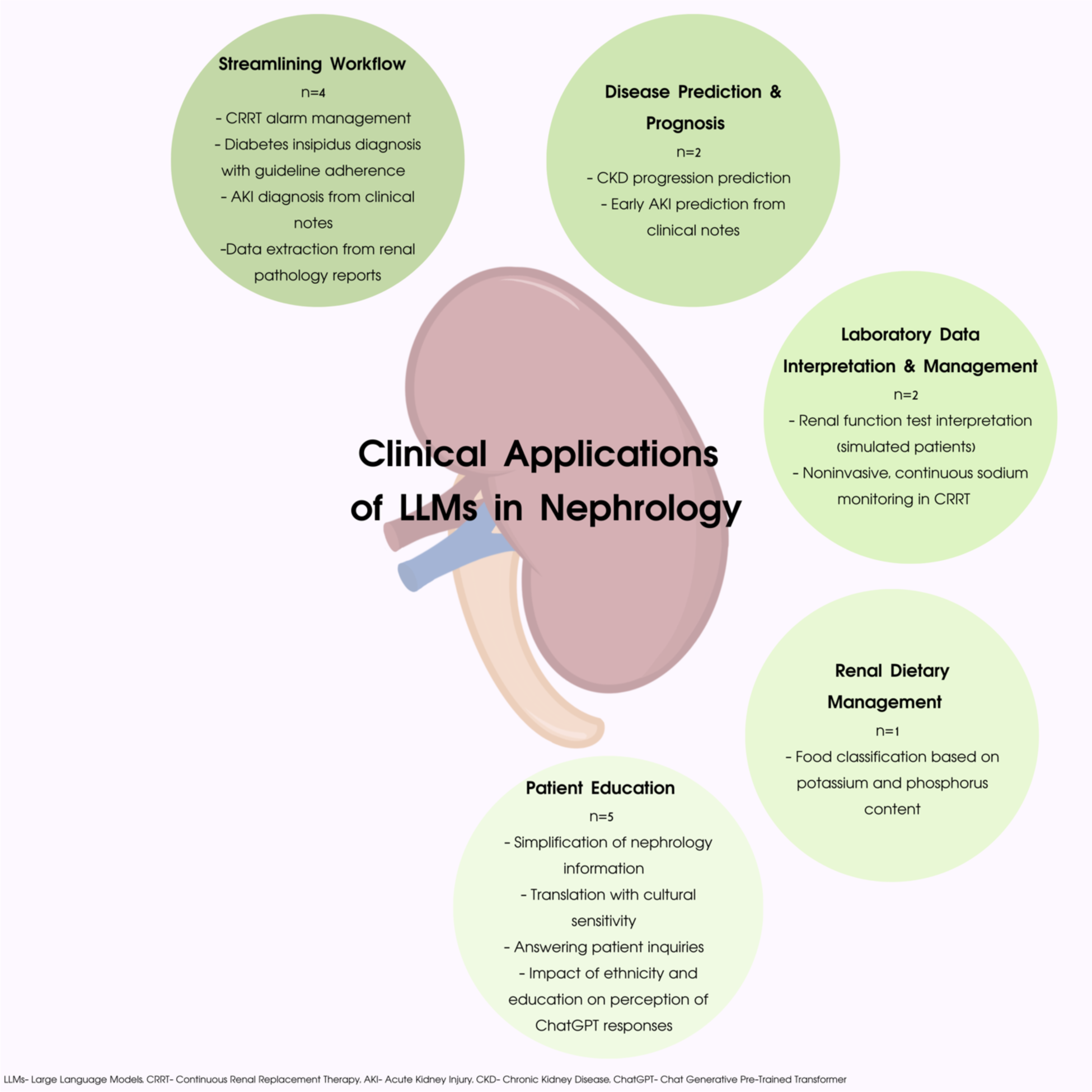
Visual Abstract.

**Figure 2:**
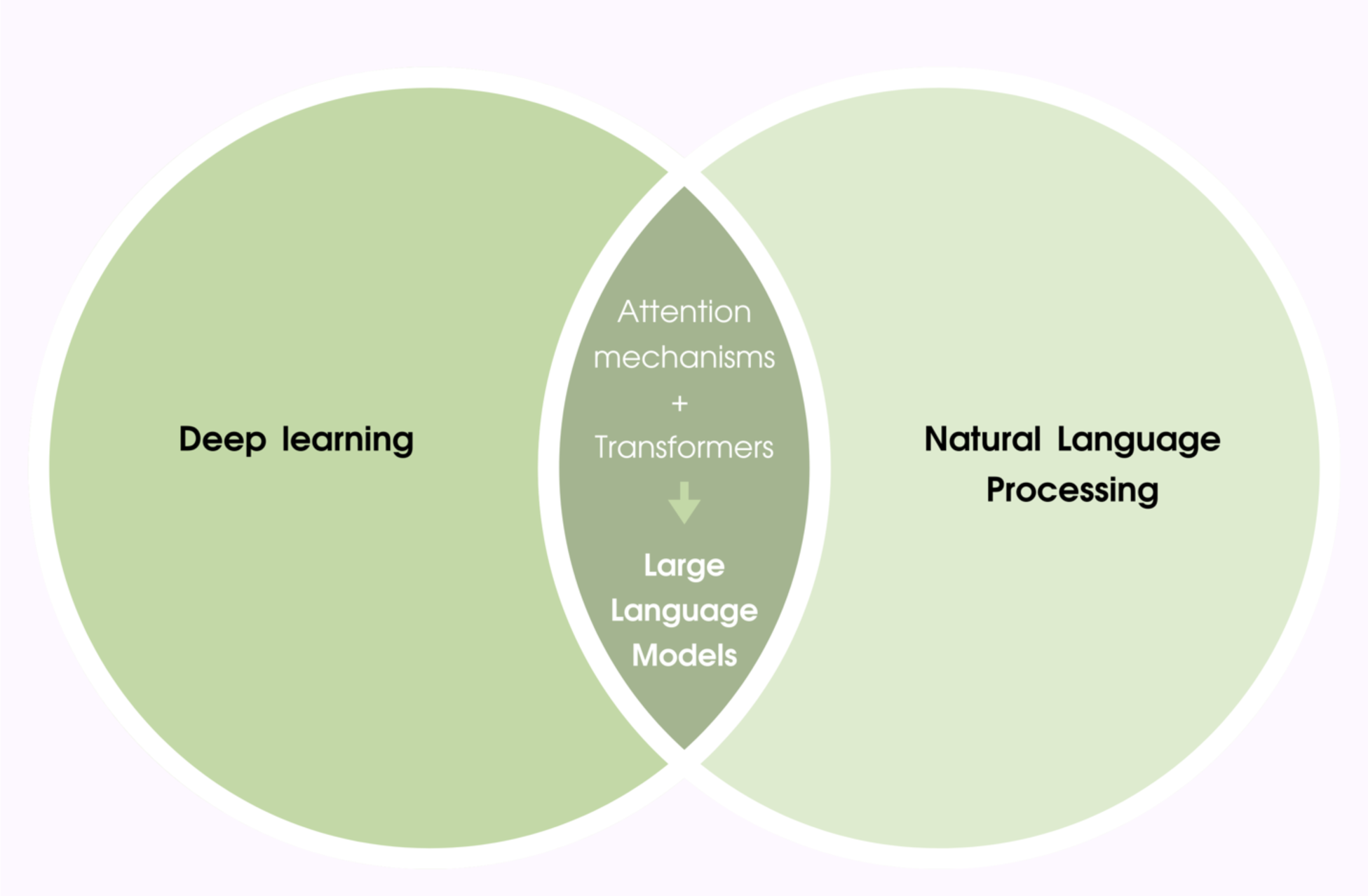
Overview of AI modalities.

*Natural Language Processing (NLP)* is an important domain within AI, offering various functions related to human language. NLP allows for human language understanding, such as human-like interactions between the user and the chatbot, text generation and processing, and many other functions ^18^.

*Deep Learning (DL)* is an advanced type of AI. Within NLP it is used to facilitate its complex linguistic functions. The underlying DL architecture is inspired by the function of biological neurons. It is based on artificial neural networks arranged in multiple layers (hence “deep”). Data processing is handled by interconnected nodes representing neurons, where each “neuron” is similar to a single logistic regression unit^19^.

When the user presents the chatbot with a textual input (termed prompt), the text then passes through several layers of interconnected nodes, each layer allowing the algorithm additional understanding of the text. An *attention mechanism* is used, detecting individual importance of words within a sentence^20^. This ultimately allows the machine to understand the written text in a contextual manner, and therefore, more accurately.

*Transformers* are a specific type of multi-layered neural network used in DL, characterized by their use of attention mechanisms. A major advancement in transformers occurred with the release of *Bidirectional Encoder Representations from Transformers (BERT)*^21^. BERT improved the application of transformer architecture and achieved state-of-the-art results in various NLP tasks. The improvement offered by BERT was due to several factors, including its bidirectional text processing ability, its pre-training and fine-tuning processes, and its effective adaptability to new tasks.

*Large Language Models (LLMs)* represent a significant development in the field of transformers and the expansion of NLP’s capabilities. LLMs enable complex language generation skills, as seen in well-known chatbots such as openAI’s Chat Generative Pre-Trained Transformer (ChatGPT)^1^ and Google’s Gemini ^22^. These platforms allow users to pose prompts, and receive written, coherent, and contextual answers generated by AI. When the prompt is more descriptive, the generated answer becomes more accurate. Language generation by LLMs is based on predicting the most probable sequence of words, one by one. LLMs are trained on very large databases and then fine-tuned via reinforcement learning—a process of improving the tool’s performance through its own experience. The powerful text analysis capabilities offered by LLMs can be widely used across numerous professions, potentially alleviating the burden of intricate data processing and information retrieval, thus enabling a more effective workflow. OpenAI offers both free and paid versions of ChatGPT, with the free version utilizing GPT-3.5 and the paid version powered by the more advanced GPT-4. In this paper, ChatGPT will refer to the GPT-3.5 version, while GPT-4 will denote the paid, more advanced model to maintain clarity between the two versions.

As AI continues to evolve, its applications in various medical fields are becoming increasingly prominent^3–11^. This systematic review aims to explore how the diverse functions of LLMs can be leveraged to enhance clinical care within the field of nephrology.

## Methods

### Search strategy

This systematic review followed the Preferred Reporting Items for Systematic Reviews and Meta-Analyses (PRISMA) guidelines **(Figure 3)**.

**Figure 3:**
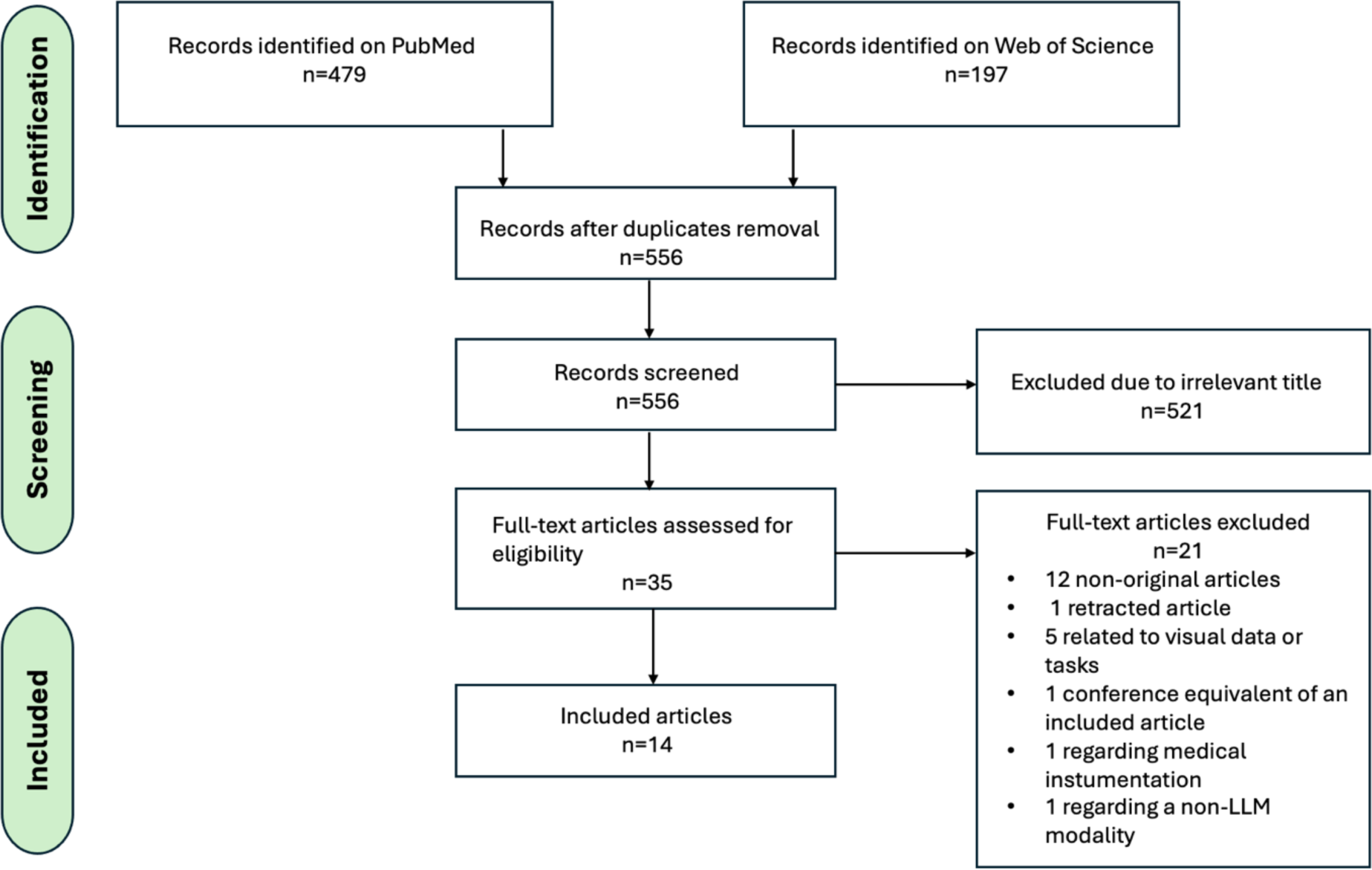
Search and selection flowchart.

We conducted a literature search in MEDLINE/PubMed and Web of Science databases on July 21^st^, 2024. The keywords used for the search were related to two main subjects: “Nephrology” and “LLM”. The full strategy search for each database is detailed in **Supplementary Material 1**.

Studies were included in this systematic review if they addressed the clinical applications of LLMs within the field of nephrology. To meet the inclusion criteria, studies had to be peer-reviewed original research articles published in English and directly relevant to the integration of LLMs in nephrology practice, including aspects such as patient care, diagnostic processes, treatment efficacy, and clinical outcomes.

Exclusion criteria were applied to ensure the relevance and quality of the review. Non-original articles, including reviews, editorials, and commentaries, were excluded. Studies that focused on areas outside of nephrology, such as urology, renal oncology, clinical pharmacology, and laboratory medicine instrumentation, were omitted. Additionally, research concentrating on LLM optimization techniques, engineering applications, or technological advancements without a clear connection to patient care or clinical outcomes in nephrology was excluded. Articles evaluating AI tools in non-clinical settings—such as professional certification assessments, literature search assistance, scientific writing support, or exam question responses—did not meet the inclusion criteria. Furthermore, studies exploring non-LLM artificial intelligence applications or those showcasing visual data processing tasks performed by LLMs were excluded.

This systematic review is registered in PROSPERO: CRD42024550169.

### Study selection

Two reviewers (ZU and SS) independently screened the titles and abstracts to decide whether the results met the inclusion criteria. A further review of the full-text article took place in case of uncertainty. A third reviewer (EK) aided in solving any disagreements in the study selection process.

### Data extraction

Data was collected using a standardized data extraction sheet. The information gathered included the year of publication, study design, study location, ethical statement, number of patients, inclusion and exclusion criteria, description of the population, use of an online database, size of the online database, use of an independent test dataset, clinical application, evaluation metrics, and performance results.

### Quality assessment and risk of bias

To check for bias and quality assessment, we used the adapted version of Quality Assessment of Diagnostic Accuracy Studies (QUADAS-2) criteria.

## Results

### Study selection and classification process

Our literature search identified a total of 556 papers. Of these, 14 studies met our inclusion criteria **Figure 3**. We categorized the eligible studies into five distinct nephrology practice clinical applications:

Streamlining workflow, disease prediction and prognosis, laboratory data interpretation and management, renal dietary management, and patient education, as outlined in **Table 1**.

**Table 1:**

| Clinical application | Title | 1<sup>st</sup> author | Journal | Year of publication |
| --- | --- | --- | --- | --- |
| Streamlining workflow | Personalized Medicine Transformed: ChatGPT's Contribution to Continuous Renal Replacement Therapy Alarm Management in Intensive Care Units | Sheikh MS<sup>23</sup> | J Pers Med | 2024 |
|  | How to improve ChatGPT performance for nephrologists: a technique guide | Miao J<sup>24</sup> | J Nephrol | 2024 |
|  | Constructing synthetic datasets with generative artificial intelligence to train large language models to classify acute renal failure from clinical notes | Litake O<sup>25</sup> | J Am Med Inform Assoc | 2024 |
|  | exKidneyBERT: a language model for kidney transplant pathology reports and the crucial role of extended vocabularies | Yang T<sup>26</sup> | PeerJ Comput Sci | 2024 |
| Disease prediction and prognosis | Transformer-based time-to-event prediction for chronic kidney disease deterioration | Zisser M<sup>29</sup> | J Am Med Inform Assoc | 2024 |
|  | A Pre-trained Clinical Language Model for Acute Kidney Injury | Mao CS<sup>30</sup> | Proceedings of the 2020 8th IEEE International Conference on Healthcare Informatics (ICHI 2020) | 2020 |
| Laboratory data interpretation and management | Response accuracy of ChatGPT 3.5 Copilot and Gemini in interpreting biochemical laboratory data a pilot study | Kaftan AN<sup>15</sup> | Sci Rep | 2024 |
|  | Contactless and continuous sodium concentration monitoring during continuous renal replacement therapy | Berger M<sup>31</sup> | Sensors and Actuators B: Chemical | 2020 |
| Renal dietary management | AI-Powered Renal Diet Support: Performance of ChatGPT, Bard AI, and Bing Chat | Qarajeh A<sup>32</sup> | Clin Pract | 2023 |
| Patient education | Empowering inclusivity: improving readability of living kidney donation information with ChatGPT | Garcia Valencia OA<sup>33</sup> | Front Digit Health | 2024 |
|  | AI-driven translations for kidney transplant equity in Hispanic populations | Garcia Valencia OA<sup>34</sup> | Sci Rep | 2024 |
|  | Using ChatGPT for Kidney Transplantation: Perceived Information Quality by Race and Education Levels | Lee J<sup>35</sup> | Clin Transplant | 2024 |
|  | Can large language models provide accurate and quality information to parents regarding chronic kidney diseases? | Naz R<sup>36</sup> | J Eval Clin Pract | 2024 |
|  | Evaluating ChatGPT's Accuracy in Responding to Patient Education Questions on Acute Kidney Injury and Continuous Renal Replacement Therapy | Sheikh MS<sup>37</sup> | Blood Purif | 2024 |

The characteristics of each included article are presented in **Table 2**. Furthermore, a thorough comparison of the advantages and limitations of the discussed LLM modalities is in **Table 3**.

**Table 2:**
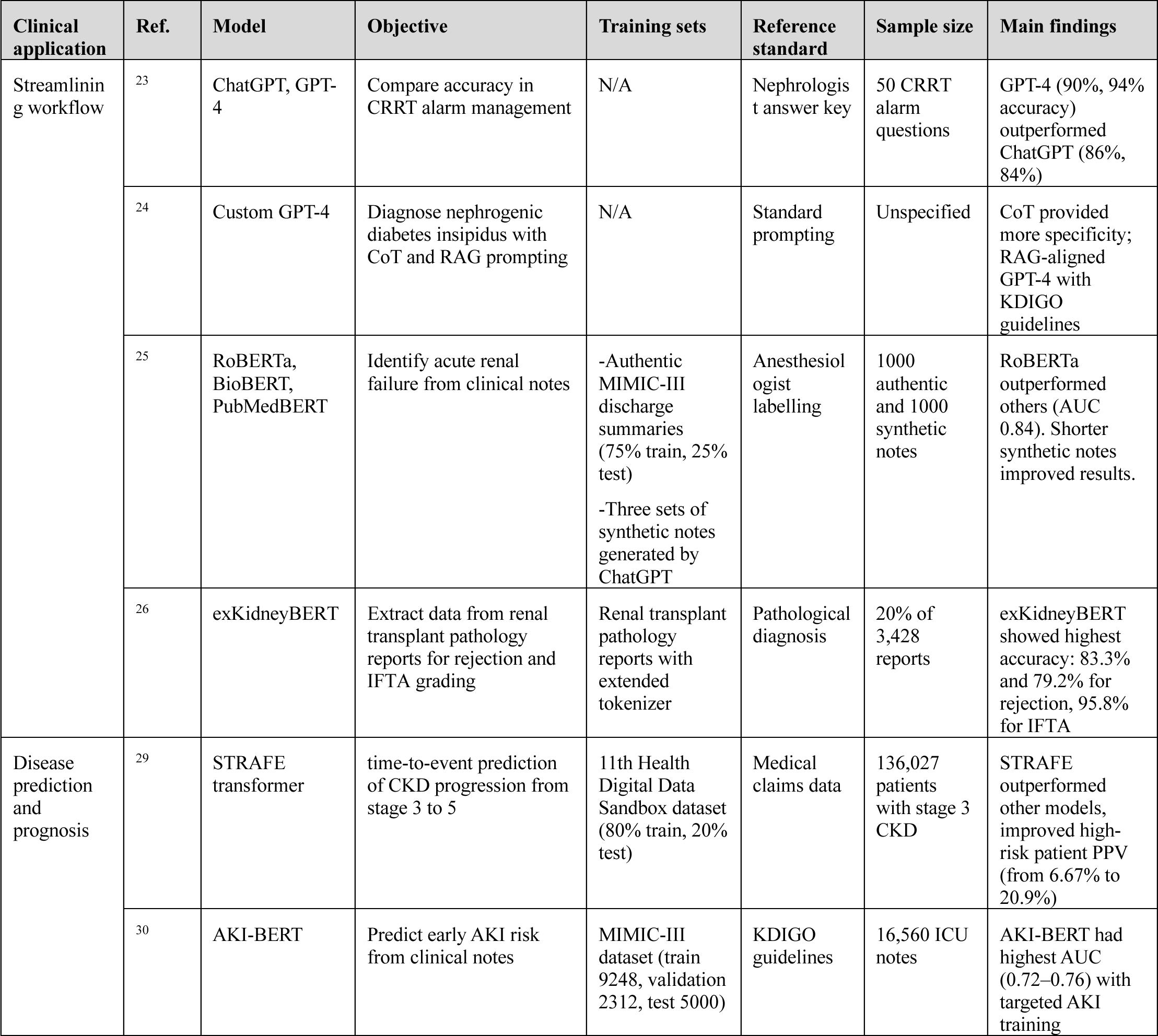
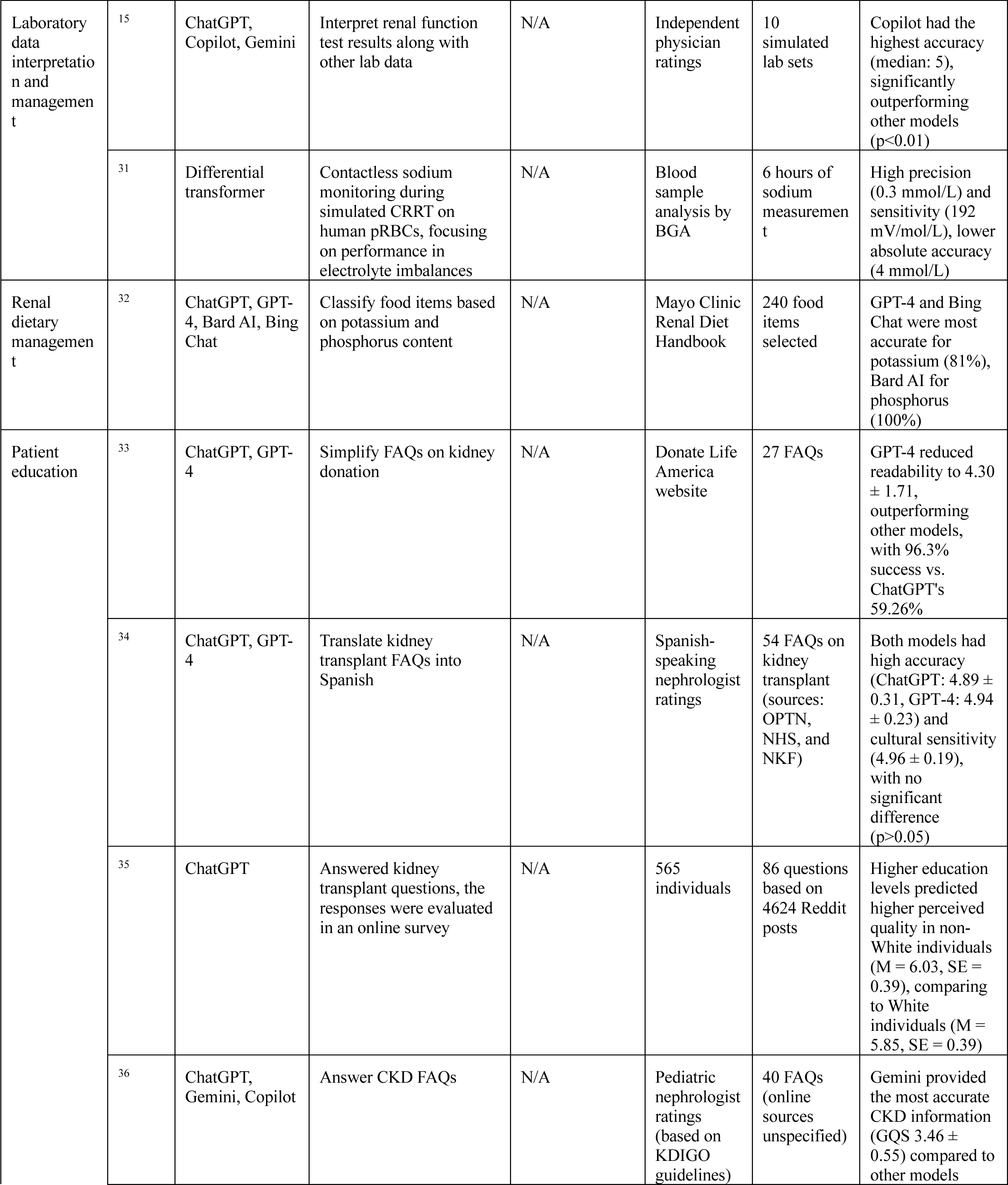
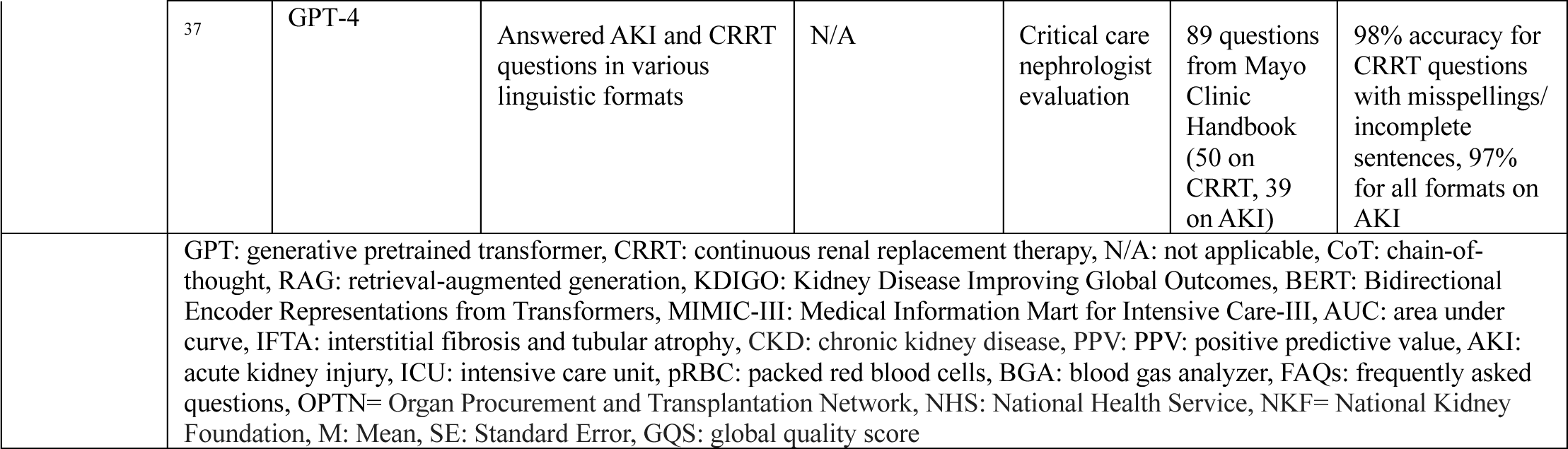

**Table 3:**
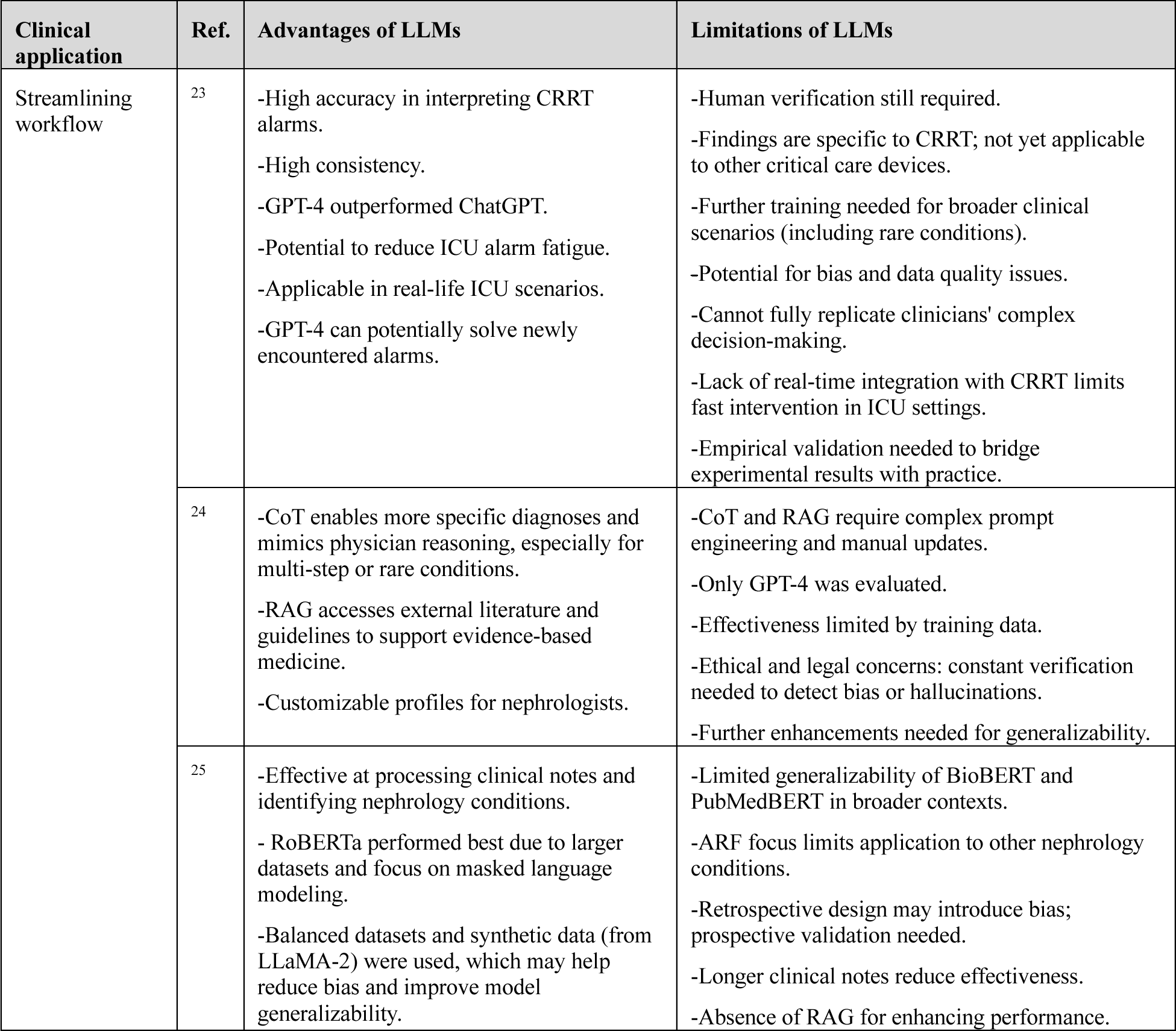
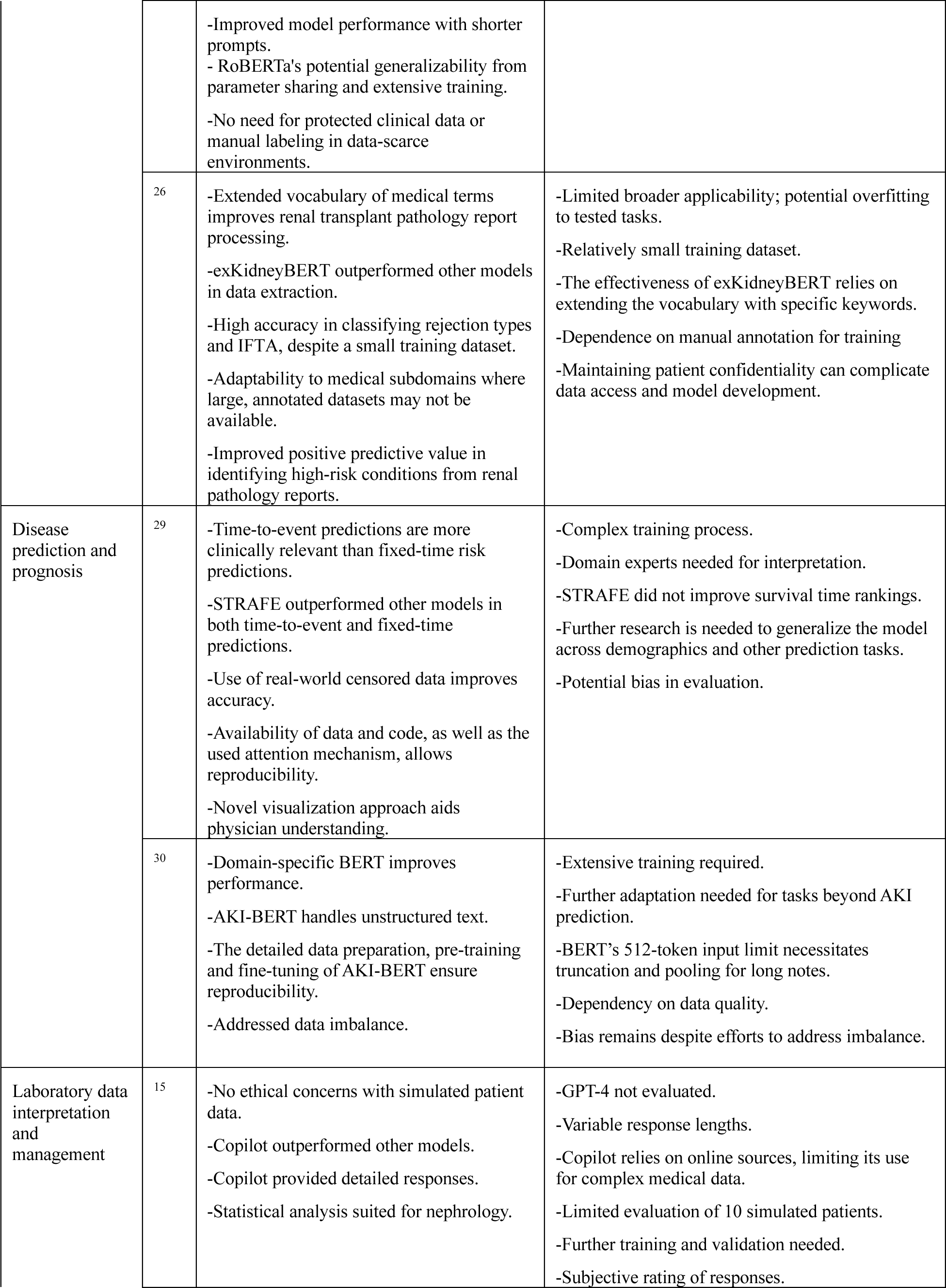
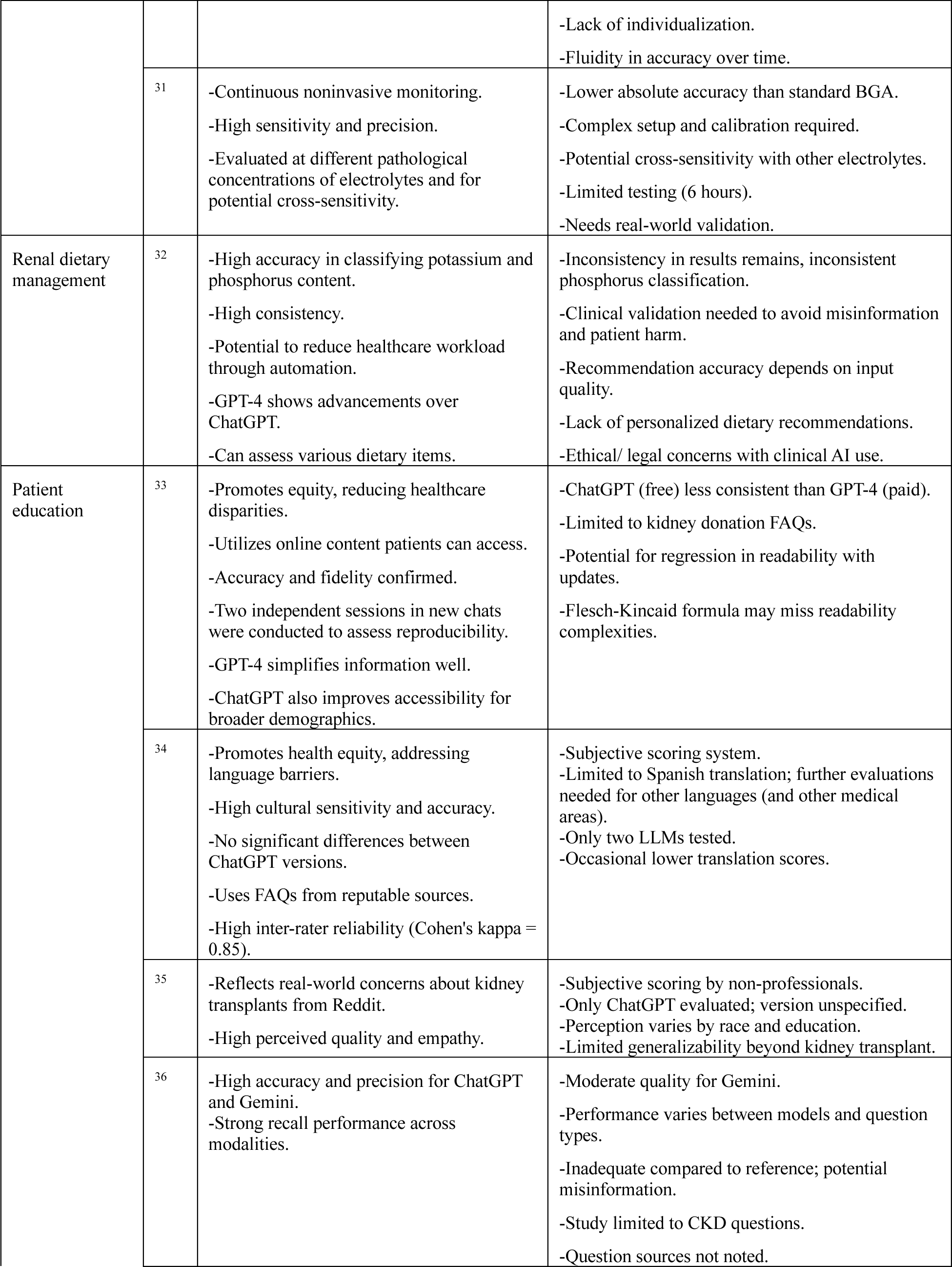
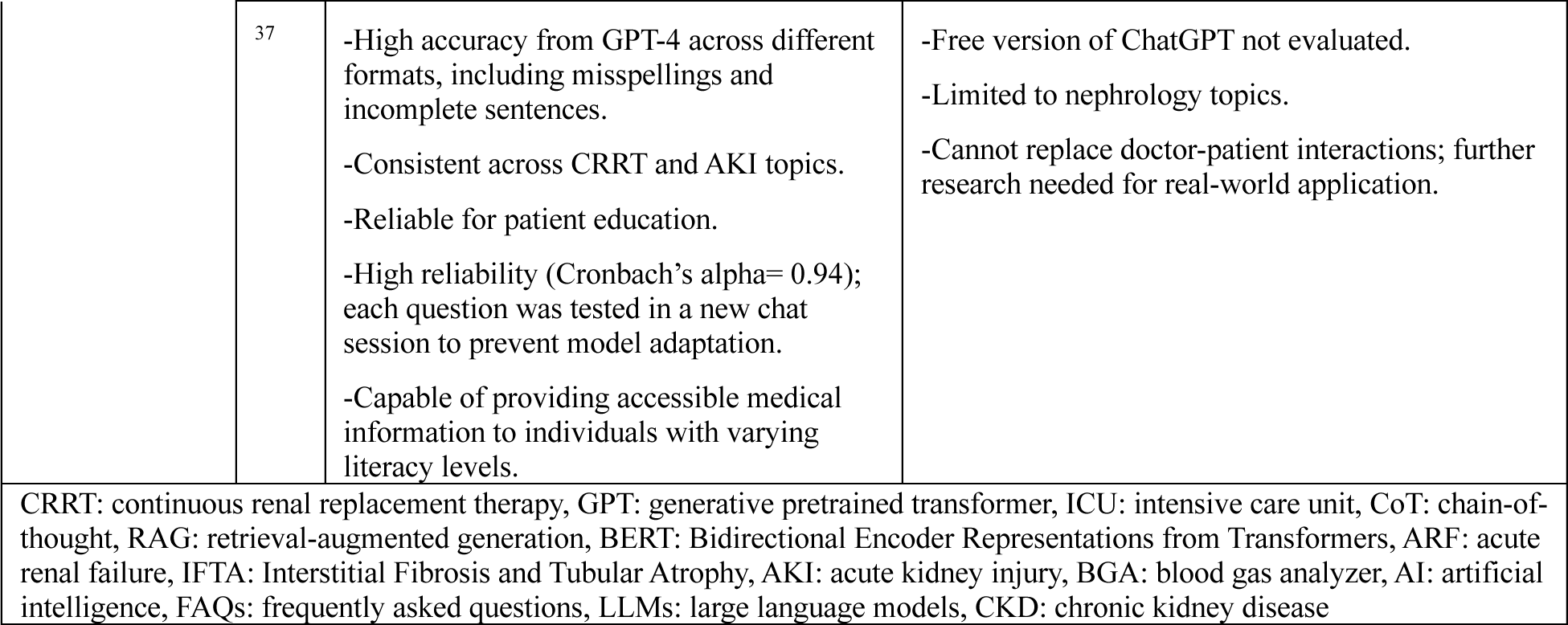

### Quality assessment

To assess the quality and risk of bias, we employed the adapted version of the Quality Assessment of Diagnostic Accuracy Studies (QUADAS-2) criteria across the 14 studies. Overall, all studies exhibited a moderate to high risk of bias concerning the index test, predominantly due to the absence of external validation and ambiguity surrounding the independence of results interpretation relative to the reference standard. Notably, two studies were classified as having a high risk of bias in at least one domain. The data management domain also revealed a moderate risk of bias in most studies, primarily due to incomplete or poorly specified data management processes and a lack of clear procedures to ensure data integrity. The detailed quality assessment is available in **Supplementary Material 2**.

### Descriptive summary of the results according to the five clinical applications

#### Streamlining workflow

Four papers were classified under this category, each addressing distinct aspects of clinical work performed by nephrologists ^23–26^

Sheikh MS et al.^23^ demonstrated high accuracy of GPT-4 (90-94%) in managing continuous renal replacement therapy (CRRT) alarms, surpassing the performance of ChatGPT (84-86%) although the difference was not statistically significant. As the chosen CRRT questions reflect real-life frequently encountered ICU scenarios, the study illustrates LLMs’ potential to reduce ICU alarm fatigue and improve patient safety.

Miao J et al.^24^ provided a descriptive application of GPT-4 in addressing inquiries from practicing nephrologists, using nephrogenic diabetes insipidus (DI) diagnosis as a case study. They employed techniques such as Chain of Thought (CoT) prompt engineering, which guides the model to break down reasoning step-by-step ^27^, and Retrieval-Augmented Generation (RAG), which integrates external data sources into the model’s responses ^28^. The authors found that these methods enhance diagnostic specificity and alignment with the *kidney disease: Improving Global Outcomes (KDIGO)* guidelines.

#### Disease prediction and prognosis

Two studies presented LLM tools for different prediction tasks related to nephrology conditions ^29,30^ Zisser M et al.^29^ introduced the STRAFE transformer, which outperformed other models in predicting progression to stage 5 chronic kidney disease (CKD) and significantly improving the identification of high-risk patients. STRAFE’s ability to utilize real-world censored data (of patients with limited observation time, who have not yet encountered the event of interest) allows a more accurate time-to-event prediction.

Mao CS et al.^30^ presented AKI-BERT for the early prediction of acute kidney injury (AKI) based on clinical notes. AKI-BERT achieved higher accuracy (AUC 0.720-0.764) compared to general BERT models, emphasizing the importance of specialized training in handling unstructured medical text. Both models require a complex training process and further adaptations for generalizability to other tasks.

#### Laboratory data interpretation and management

Two papers explored the integration of LLMs in the laboratory aspect of nephrology ^15,31^

Kaftan AN et al. ^15^compared the performance of three LLMs in interpreting ten simulated sets of laboratory values, and found that the Copilot model demonstrated the highest accuracy. Copilot’s performance had statistically significant difference from ChatGPT’s (p=0.002 for all lab results, 0.001 for kidney functions) and Gemini’s (p=0.008 for all lab results, 0.005 for kidney functions). It is noteworthy that the newer GPT-4 model was not evaluated, and only ten cases were analyzed. Despite the high performance of Copilot, it relies on input quality and bases its responses on online resources, which are not necessarily tailored for professional medical use.

Berger M et al. ^31^introduced a differential transformer for sodium monitoring during a simulated CRRT setup, utilizing a noninvasive and contactless architecture. The test duration was limited to six hours, during which electrolytes were intentionally varied to assess performance in pathological states as well, which are commonly encountered in patients requiring CRRT.

The transformer demonstrated high sensitivity to sodium concentration changes (192 mV/mol/L) and greater precision in repeated measurements (0.3 mmol/L) compared to standard blood gas analyzer (BGA), which has a precision of 0.6 mmol/L. While the absolute accuracy was 4 mmol/L, lower than the BGA’s 2 mmol/L, this was still considered sufficient for continuous sodium monitoring. Further evaluation of this tool in real-world clinical settings is needed.

#### Renal dietary management

The renal diet, an essential part of care for patients with kidney disease, was addressed by one study ^32^

Qarajeh A et al. evaluated the ability of four different LLMs to classify food based on their potassium and phosphorous content. GPT-4 and Bing Chat excelled in potassium content classification, achieving an accuracy of 81%, while Bard AI showed 100% accuracy in determining phosphorus content.

#### Patient education

The abilities of LLMs to answer patient inquiries and concerns were showcased in five papers ^33–37^

Garcia Valencia OA et al. ^33,34^contributed two significant studies to this group, both promoting inclusivity and equity in renal healthcare. The first paper ^33^assessed the simplification abilities of ChatGPT and GPT-4 for 27 frequently asked questions (FAQs) and answers on kidney donation. Two independent attempts were conducted in new chat sessions. GPT-4 significantly reduced the average reading grade level from the original reference of 9.6 ± 1.9 (roughly 10^th^ grade) to 4.30 ± 1.71 (4^th^ grade), while ChatGPT reduced it to 7.72 ± 1.85 (about 8^th^ grade). The goal was to simplify the text to below an 8^th^-grade reading level. GPT-4 achieved this in 96.30% of cases, while ChatGPT succeeded in 59.26% of cases, indicating that the free version of ChatGPT provides more limited access to high-quality, simplified information. The second paper by this author ^34^, evaluated the translation abilities from English to Spanish of 54 FAQs on kidney transplant. Two Spanish-speaking nephrologists scored the translations and found high levels of linguistic accuracy (ChatGPT: 4.89 ± 0.31, GPT-4: 4.94 ± 0.23) and cultural sensitivity for Hispanics (4.96 ± 0.19 for both models) in both ChatGPT and GPT-4. As opposed to the previous paper, there was no significant difference between the performance of the paid and free versions of ChatGPT (linguistic accuracy: p=0.26, cultural sensitivity: p=1.00).

Lee J et al.^35^ also evaluated 86 questions on kidney transplant, selected from a pool of real questions asked on Reddit. In this research, the rating of information quality and empathy was done by 565 participants in an online survey. While the study did not explicitly exclude medical professionals as raters, the primary aim was to capture individual perceptions of ChatGPT-generated responses, reflecting how they might be received by patients. The study found that higher education levels among non-White individuals predicted higher-perceived quality (M = 6.03, SE = 0.39), whereas higher education among White individuals led to lower perceived quality (M = 5.85, SE = 0.39).

Similarly, Naz R et al. ^36^ researched the accuracy and quality of information provided by three LLMs, when asked 40 FAQs on a different topic-parents’ concerns about CKD. Two independent pediatric nephrologists classified the generated responses with respect to the KDIGO guidelines as a reference. ChatGPT and Gemini showed high accuracy in diagnosis and CKD lifestyle questions. Among the models evaluated, Gemini was noted as the most accurate in providing information on CKD, with an average Global Quality Score (GQS) of 3.46 ± 0.55.

## Discussion

In this systematic review we explored the use of various LLMs such as ChatGPT in nephrology. We focused on five key aspects of patient care emphasizing their potential to enhance both physician workflows and patient engagement. However, the tested modalities present several limitations, including dependence on input quality, and the necessity for further validation in diverse clinical settings.

### Applications from Physicians’ Perspective

LLMs, such as GPT-4, have demonstrated significant potential in improving workflow efficiency in nephrology, particularly in ICU settings. However, several of these tools, including their application in continuous renal replacement therapy (CRRT) management, lack extensive external validation and have not been prospectively tested in real-world clinical environments ^2331^. Additionally, LLMs support diagnostic processes by enhancing diagnostic specificity and ensuring alignment with established guidelines like KDIGO, thereby improving clinical decision-making and patient outcomes ^24^To mitigate issues such as hallucinations and outdated information, Retrieval-Augmented Generation (RAG) has been integrated to pull from external data sources, ensuring that LLMs provide up-to-date, guideline-adherent recommendations ^24,28^. However, constant verification is required, and ethical issues related to cloud-based patient data processing and security pose significant barriers to widespread implementation.

LLMs also contribute to disease prediction by accurately forecasting CKD progression and AKI, facilitating early interventions. Models like STRAFE and AKI-BERT leverage unstructured clinical data to identify high-risk patients, enhancing personalized patient management. In laboratory data interpretation, LLMs improve the accuracy of renal function test analyses.

While these developments show promise, the ability of tools like ChatGPT to simulate a physician’s thought process remains limited. Although LLM reasoning mimics a physician’s diagnostic approach by breaking the reasoning process into steps, these models still require further validation before being fully integrated into clinical practice^24,27^. For instance, Kaftan AN et al. showcased Copilot‘s accuracy in interpreting ten sets of laboratory values; however, its reliance on online resources and its inability to manage complex medical data suggest that further empirical validation in real-world settings is needed ^15^. Moreover, despite their potential, the generalizability of these models remains restricted, necessitating further validation before widespread adoption in nephrology practice.

### Applications from Patients’ Perspective

The incorporation of LLMs into nephrology practice holds the potential for bridging gaps in patient education and improving accessibility to medical information. For example, models such as GPT-4, can simplify complex medical concepts, provide culturally sensitive translations, and accurately respond to frequently asked questions, even when inquiries contain misspellings or are incomplete. ^33,34,37^. The studies reviewed focused on patient inquiries based on real online sources, that patients may encounter when exploring a nephrology subject online. These studies underscore the potential for AI tools to improve the accessibility of health-related content across different literacy levels and languages, promoting inclusivity and health equity ^33,34^. However, a significant disparity remains between the free and paid versions of ChatGPT, with GPT-4 (the paid version) consistently outperforming the free version in medical information simplification tasks ^33^. This performance gap, while showcasing the advancements in AI, also raises concerns about accessibility, as better-quality health information is currently limited to those who can afford paid access, highlighting an inherent health inequity.

While LLMs enhance accessibility, they may oversimplify information, potentially omitting critical details necessary for comprehensive patient understanding ^38^. Furthermore, as Lee J et al. noted, while tailoring AI-generated responses based on education level and race has the potential to improve effectiveness, patient perception of these responses can still vary, suggesting that LLM-generated information may not always be equally accessible or comprehensible to all patients ^35^. Lastly, the lack of human interaction in LLMs limits their ability to provide empathetic, personalized care, a crucial aspect of effective doctor-patient communication ^37^. This limitation raises ethical concerns about their integration into clinical settings.

### Limitations

Our systematic review focused on five areas of possible clinical applications of LLMs in nephrology, specifically excluding non-clinical applications. Some of the papers retrieved relied on a relatively small dataset. The heterogenicity in the tested clinical tasks and used methods among our selected papers did not allow us to conduct a meta-analysis. The field of LLMs and its clinical implications is quickly evolving, and thus drawing conclusions still requires further research.

In conclusion, while incorporating LLMs in nephrology shows promise across various levels of patient care, their broad implementation is still premature. Further research is required to validate these tools in terms of accuracy, rare and critical conditions, and real-world performance.

## Contributions

ZU: Data collection and extraction, writing the manuscript, interpretation and visualization of the presented results.

SS: Provided critical guidance throughout, contributed to data interpretation, and thoroughly revised the manuscript.

OE: Provided critical guidance throughout, contributed to data interpretation, and thoroughly revised the manuscript.

LC: Provided critical guidance throughout, contributed to data interpretation, and thoroughly revised the manuscript.

EK: Provided critical guidance throughout, contributed to data interpretation, and thoroughly revised the manuscript.

GNN: Provided critical guidance throughout, contributed to data interpretation, and thoroughly revised the manuscript.

## Data Availability

As this article is a systematic review, no new data was generated. Instead, previously published data were analyzed. All data related to this systematic review is included in this published article and its supplementary materials. Any additional data are available upon reasonable request to the authors.

