## Supplementary Materials for "Clinical Applications and Limitations of Large Language Models in Nephrology: A Systematic Review"

**Table of Contents:**

Supplementary Material 1- Literature Search…………..……………………………………...……….2

1.A) PubMed Search

1.B) Web of Science Search

Supplementary Material 2- Quality Assessment of Diagnostic Accuracy Studies (QUADAS-2)……...3

1. Literature Search

1.A) Pubmed Search:

**((((((((((((((((((((((((((kidney*) OR (renal)) OR (nephrology)) OR (kidney disease*)) OR (renal disease*)) OR (renal disorder*)) OR (nephropath*)) OR (acute kidney injury)) OR (acute renal failure)) OR (AKI)) OR (acute renal insufficiency)) OR (chronic kidney disease)) OR (CKD)) OR (chronic renal failure)) OR (chronic renal insufficiency)) OR (hemodialysis)) OR (dialysis)) OR (renal replacement therapy)) OR (glomerulonephritis)) OR (glomerulopath*)) OR (glomerular disease*)) OR (glomerular disorder*)) OR (nephrotic syndrome)) OR (nephritic syndrome)) OR (urinalysis)) OR (renal function test*)) AND ((((((((((large language model*) OR (LLM)) OR (ChatGPT)) OR (OpenAI)) OR (Microsoft bing)) OR (Google bard)) OR (Google gemini))) OR (BERT)) OR (transformer*))**

**1.B) Web of Science Search:**

TS=((kidney* OR renal OR nephrology OR "kidney disease*" OR "renal disease*" OR "renal disorder*" OR nephropath* OR "acute kidney injury" OR "acute renal failure" OR AKI OR "acute renal insufficiency" OR "chronic kidney disease" OR CKD OR "chronic renal failure" OR "chronic renal insufficiency" OR hemodialysis OR dialysis OR "renal replacement therapy" OR glomerulonephritis OR glomerulopath* OR "glomerular disease*" OR "glomerular disorder*" OR "nephrotic syndrome" OR "nephritic syndrome" OR urinalysis OR "renal function test*") AND (("large language model*" OR LLM OR ChatGPT OR OpenAI OR "Microsoft bing" OR "Google bard" OR "Google gemini" OR BERT OR transformer*)))

2. Quality Assessment of Diagnostic Accuracy Studies (QUADAS-2)

Index:

N/A= not applicable

V= low risk of bias

X= high risk of bias

M=moderate risk of bias

| Author / risk of bias | Patient selection | Index test | Reference standard | Flow and timing | Data management |
| --- | --- | --- | --- | --- | --- |
| Sheikh MS et al. | N/A | X* | V** | N/A | V |
| Miao J et al. | N/A | M* | M | N/A | V |
| Litake O et al. | N/A | M* | V | N/A | M**** |
| Yang T | N/A | M | V | N/A | V |
| Zisser M et al. | V | M | V | V | M**** |
| Mao CS et al. | V | M | V | V | V |
| Kaftan AN et.al | X | M | V | V | X |
| Berger M et al. | N/A | M* | V | V | M |
| Qarajeh A et al. | N/A | M | V | N/A | M |
| Garcia Valencia OA et al. | N/A | M* | V | N/A | M |
| Garcia Valencia OA et al. | N/A | M* | V | N/A | M |
| Lee J et al. | M*** | M* | M | N/A | M |
| Naz R et al. | N/A | M* | V | N/A | M |
| Sheikh MS et al. | N/A | M* | M | N/A | M |

* Of note, the setting is experimental

**Not universally accepted

***Voluntary online survey

****Conflict of interest
